# LINK WORKER LED SOCIAL PRESCRIBING AS EARLY INTERVENTION FOR CHILDREN WITH SOCIAL-EMOTIONAL MENTAL HEALTH DIFFICULTIES

**DOI:** 10.1101/2024.12.17.24319112

**Authors:** J. J. V Charlton, A Battersby, C Drinkwater, M McKean, T Quibell

## Abstract

**Objective:** To evaluate the impact of a novel approach to Social Prescribing (SP) embedded within education for primary-school aged children with early social-emotional mental health (SEMH) difficulties in socioeconomically deprived areas

**Design:** Repeated measures

**Setting:** 10 primary schools

**Patients:** 117 children aged 7-11 years with SEMH

**Intervention:** Children received weekly support with a Link Worker and support to attend community assets over 9 months.

**Main outcome measures:** Teacher (T) and parent (P) report Strengths and Difficulties Questionnaire measured SEMH, parent and child report Paediatric Quality of Life Inventory measured quality of life, and a child report vocabulary list measured expressive and receptive vocabulary.

**Results:** There was a significant effect of provision on SEMH reported by teachers (n-71) and parents (n=92), including total difficulties (95% CI .000-.001; 95% CI .015-.020), conduct difficulties (95% CI .012-.017; 95% CI .000-.001) and hyperactivity (95% CI .000-.001; 95% CI .000-.000, respectively) with moderate effect sizes. Parents (n=74) reported a significant improvement in children’s emotional (95% CI .066-.075) and school functioning (95% CI .005-008) with small effect sizes, however child reported quality of life (n=83) remained stable. There was a significant improvement in children’s receptive vocabulary (95% CI .019-.025) with moderate effect size, but not expressive vocabulary.

**Conclusions:** Outcomes indicate a potential role for SP embedded within education as a preventative strategy in children’s early social-emotional mental health and wellbeing. Further work is required to explore direct impacts on mental health services and long-term impacts on SEMH.

**What is already known on this topic:** Child-centred SP is emerging as an alternative approach to support within health and voluntary sectors, however there is a gap in SP embedded within education, and limited evidence for the impact of SP on child SEMH and wellbeing.

**What this study adds:** Early evidence for education-embedded SP as a potential preventative strategy in children’s mental health and wellbeing.

**How this study might affect research, practice or policy:** Outcomes have potential implications for supporting capacity of primary and secondary care services, a framework for collaborative delivery of support between education, health and voluntary sector services, and a defined SP model for further research.

## INTRODUCTION

The sharp rise in presentations of children with primary mental health problems to paediatric services reflects the tip of the iceberg of this overwhelming public health crisis. Childhood mental health disorders in England have increased significantly over the past decade; an additional 500,000 children and young people experience mental health problems compared to 2017, 20.3% of which are children aged 8-16 years^1^. Social-emotional mental health (SEMH) difficulties are the second most common type of Special Educational Need (SEN) at 22.3%, behind speech, language and communication needs (SLCN) at 25.6%^2^, both of which have profound long-term impacts for development and life opportunities^3,4^. Most at-risk are children experiencing poverty^5^; there is a stark gap between the wellbeing of children living in poorer areas and those in more affluent areas.

Traditional mental health services are overwhelmed, with lengthy waits for input potentially delaying intervention and resulting in an escalation of problems. It is estimated that over a quarter of referrals to Children and Young People’s Services for children from GPs are rejected due to conditions being unsuitable for treatment or not meeting eligibility criteria, and there are waiting times for secondary services of up to 6 months^6^. Yet childhood is also a critical developmental period and a period in which there is an opportunity for children to develop resilience strategies, and reduce pressure on public health services, in the future.

NHS England has supported Social Prescribing (SP) as one of its 10 high impact actions for primary care^7^. SP seeks to address healthcare inequalities, bridging the gap between clinical and non-clinical practice^8^ and is being increasingly utilised in primary care for adults with promising outcomes^9^. The approach involves ‘active signposting’ to integrate patients into community-based resources or ‘assets’, supported by a non-clinical Link Worker (LW), and advocates for a personalised approach to healthcare and more effective partnerships between patients and professionals^10^. The success of these interventions can be seen through reduced or more appropriate healthcare attendances and improvement in physical and mental health and wellbeing.

The increase in child mental health disorders poses a question as to whether SP may be applied in the context of early identification and intervention to improve children’s mental health outcomes, and there is indicative evidence of promise for supporting children with early mental health difficulties^11, 12^ and neuro-disability^13, 14^. Defining success can be more challenging in the paediatric population, however in the shorter term this includes impact on social and emotional wellbeing. Two key mechanisms have been suggested which may underpin SP in young people: social connectedness and behavioural activation^12^. The SP approach may be particularly useful for supporting children experiencing poverty; more than 1 in 4 children with SEMH have a parent who is unable to afford for them to attend activities outside of school^1^, which may offer therapeutic benefit to their wellbeing.

Although not as well established as adult-SP, child-centred SP has begun to change the behaviour of GPs and voluntary/statutory sector providers^9^. However there is a limited evidence-base for child SP and no established ‘gold-standard’ models for delivery. Published evidence for SP with younger samples still include those over the age of 16 rather than children^15, 12^. Systematic reviews have found that despite a wealth of eligible studies, evidence for the role of SP in improving the mental health and/or wellbeing of children is starkly lacking, fails to address developmental processes, is not underpinned by theory, and fails to include adequate outcome assessments ^16, 17^. Furthermore, despite its rise in popularity, there remains a range of definitions of SP as well as a range of intended outcomes^8^, varying within and across countries; a challenge for policy, practice, robust research and public awareness^18^. Alongside this are flexible definitions of what constitutes the role of a LW, making direct comparison between interventions challenging. Finally research indicates education programmes on average have moderate effects on child outcomes despite potential multiple confounding influences ^19^, however it remains unclear as to what the impacts of SP are when embedded within the education sector.

### Objective

The study aimed to explore the impact of a novel approach to Social Prescribing (SP) embedded within education (‘Zone West’) on the SEMH, quality of life and language skills of primary-school children with SEMH difficulties living in socioeconomically deprived communities.

## METHODS

### Design

The study was a within-subjects repeated measures design.

### Setting

The programme was delivered in 10 primary schools in Newcastle which served the 20% most deprived families, over the course of the 2022-2023 academic year. Recruitment occurred in September, and pre- and post-data were collected before children began receiving support (September/October) and after 9 months of support (June/July).

### Participants

115 children aged 7-11 years old were recruited to the study, identified for inclusion based on screening of SEMH difficulties using a standardised questionnaire completed by teachers for all children in the class; the Strengths and Difficulties Questionnaire (SDQ)^20^. Eligible children were those who had an SDQ score that fell into the ‘raised’, ‘high’, or ‘very high’ categories for degree of SEMH difficulties, and those who were not receiving any other therapeutic support. SDQ scores were considered alongside a discussion with the class teacher and Special Educational Needs Co-Ordinator (SENCO) to identify up to 15 children per school appropriate for the programme, considering any existing support in place and family circumstances. Identification primarily focused on SDQ score however consideration was also given to children for whom SEMH concerns were raised from school staff. Each recruited child was assigned a LW; there were 8 Link Workers working across the schools.

### Procedure

The Zone West Warrior Programme was guided by the ZW Theory of Change^21^, a Warrior Developmental Plan that tracked individual progress, and the Zones of Development framework (See Supplemental Material).

The programme involved children receiving weekly group-based therapeutic support with their LW in school (6 children per group) and 1-1 bi-weekly mentoring sessions which provided targeted support for individualised needs. Examples of intervention activities include social skills training, vocabulary enrichment, talking circle protocols^22^, and solution-focused approaches to problem-solving and behaviour change. Additionally, outside of school the LW linked children into established community resources or ‘assets’ that were matched to their individual developmental goals. On average, each child attended four sessions at community assets per month.

### Outcome measures

All outcome measures were completed at two time-points: pre- and post-9 months of Zone West support.

Social-emotional mental health was measured using the Strengths and Difficulties Questionnaire (SDQ)^20^, a standardised report measure of SEMH, measuring emotional difficulties, conduct difficulties, hyperactivity, peer difficulties and prosocial behaviour. Each subscale is scored out of 10 and the first four are summed to provide a total difficulties score out of 40. For all scales apart from the prosocial scale, a higher score indicates greater difficulty. The SDQ was completed by teachers and parents/carers.

### Quality of life

Quality of life (QoL) was measured using the Paediatric Quality of Life Inventory V.4.0 (PedsQL)^23^, a health status assessment that measures QoL across five functions (physical, emotional, social, and school functioning). Higher scores indicate poorer QoL in these areas. The PedsQL was completed by parents and Zone West children.

Children’s expressive and receptive vocabulary skills were measured using an author-designed vocabulary list. The list included 45 words with variable degrees of frequency expected for children aged 7-11 years. Children were asked to report how many words they understood, and how many they used. Higher scores indicated greater vocabulary skills.

The completion of child-report questionnaires was supported by LWs which facilitated trust and supported completion rate. This approach, however, presented a risk of bias in data collection. To mitigate this LWs were informed of this bias and directed to allow the children to complete the questionnaire independently and encourage ‘truthful’ and ‘honest’ responses.

### Data Analysis

Target sample size was 70 to achieve 90% power with alpha level of 0.05 and effect size 0.4 (Cohen’s d), following published guidelines^24^. Data were tested for normality distribution using the Kolmogorov–Smirnov (KS) test and found to be abnormally distributed (KS<0.05), therefore data were analysed using Wilcoxon related samples test on SPSS V27. Only matched pre-post data was included in analysis; we report the denominator for each outcome.

## RESULTS

### Demographic characteristics

Table 1 displays the demographic characteristics of the sample.

**Table 1.**
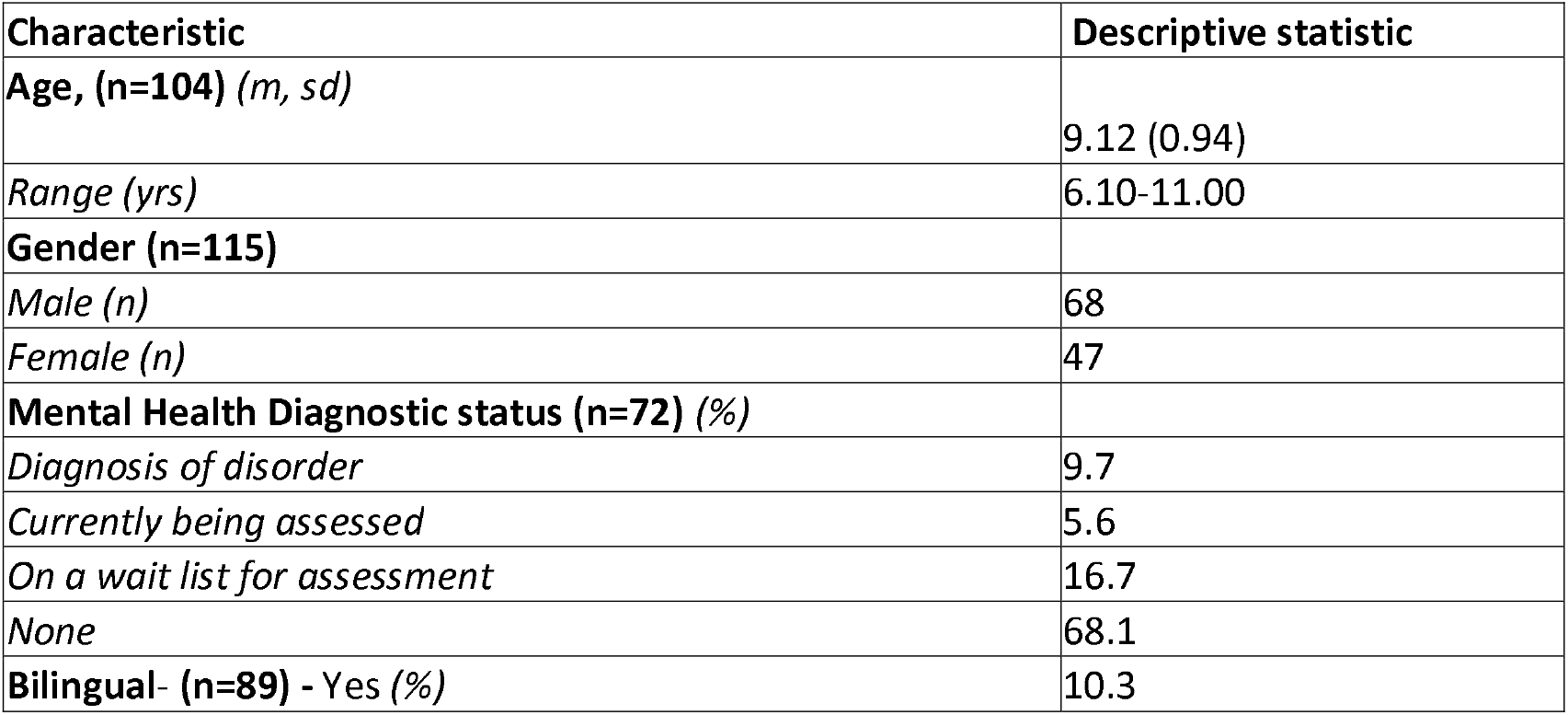
Demographic characteristics of Zone West children.

### Social-emotional mental health

Descriptive statistics for SDQ scores are displayed in Table 2.

**Table 2.**
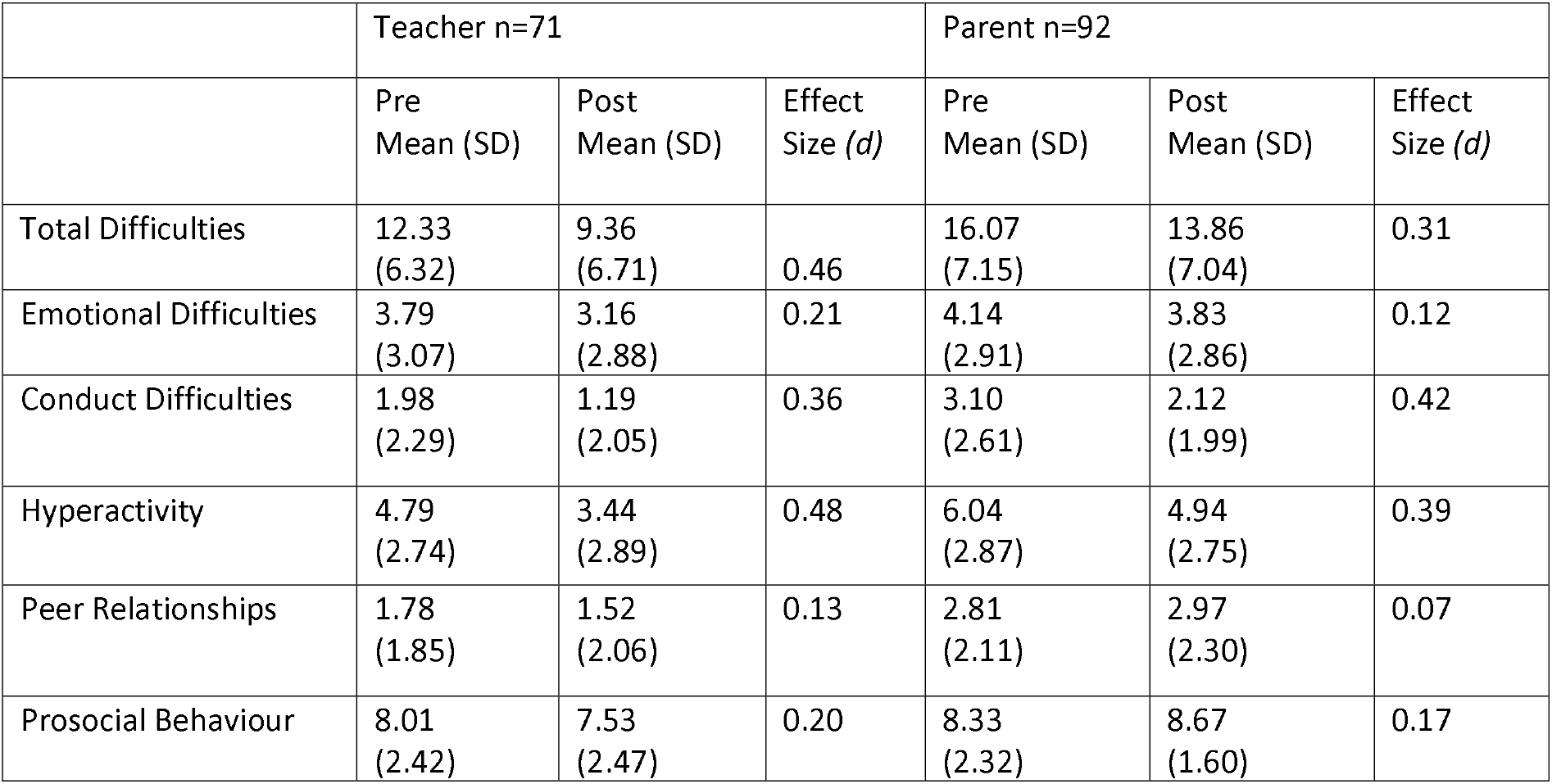
Descriptive statistics for pre- and post-teacher and parent report SDQ scores.

Based on teacher report, there was a highly significant effect of Zone West provision on children’s total behaviour difficulties (Z=−3.355, 95% CI .000-.001, p=0.001) and hyperactivity (Z=−3.290, 95% CI .000-.001, p=0.001), and a significant effect on conduct difficulties (Z=−2.442, 95% CI .012-.017, p=0.015) with moderate effect sizes, indicating a significant reduction in the degree of difficulty in these areas. There were no significant effects on emotional difficulties (Z=−1.855, 95% CI .062-0.72, p=0.064), peer difficulties (Z=−1.385, 95% CI .161-.175, p=0.166), or prosocial behaviour (Z=−0.949, 95% CI .330-.349, p=0.342).

Parent report indicated a significant effect of provision on children’s total difficulties (Z=−2.392, 95% CI .015-.020, p=0.017), and a highly significant effect on hyperactivity (Z=−3.614, 95% CI .000-.000, p=0.000) and conduct difficulties (Z=−3.942, 95% CI .000-.001, p=0.000) again with moderate effect sizes. No significant effects based on parent report were found on peer difficulties (Z=−1.164, 95% CI .233-.250, p=0.244), emotional difficulties (Z=−0.469, 95% CI .646-.645, p=0.639), or prosocial behaviour (Z=−1.567, 95% CI .89-.100, p=0.097).

### Quality of Life

Descriptive statistics for parent and child-report PedsQL scores are displayed in Table 3.

**Table 3.** Descriptive statistics for pre- and post-parent and child report PedsQL scores.

|  | Parent n=94 |  |  | Child n=94 |  |  |
| --- | --- | --- | --- | --- | --- | --- |
|  | Pre | Post | Effect Size | Pre | Post | Effect Size |

|  | Mean (SD) | Mean (SD) | (d) | Mean (SD) | Mean (SD) | (d) |
| --- | --- | --- | --- | --- | --- | --- |
| Physical | 2.07 (3.46) | 1.95 (3.46) | 0.36 | 4.10 (3.39) | 4.31 (3.94) | 0.06 |
| Emotional | 6.58 (3.94) | 6.03 (3.71) | 0.14 | 5.80 (3.23) | 6.02 (3.97) | 0.06 |
| Social | 3.46 (2.96) | 2.76 (3.14) | 0.23 | 2.92 (2.69) | 3.03 (2.85) | 0.04 |
| School | 4.70 (3.77) | 3.84 (3.53) | 0.24 | 5.33 (2.94) | 5.77 (3.32) | 0.14 |

Based on parent-report there was a significant improvement on children’s overall emotional functioning (Z= −.460, 95% CI .066-.075, p<0.05), and school functioning (Z=−2.311, 95% CI .005-008, p<0.05) with small effect sizes. Physical and social functioning remaining stable over time with no significant change (Z=−0.035, 95% CI .395-.414, p=0.97; Z=−1.008, 95% CI .111-.124, p=0.31, respectively). Based on child-report quality of life across all 4 domains remained stable with no significant changes over time; physical (Z=−0.006, 95% CI .489-.509, p=0.996), emotional (Z=−0.460, 95% CI .399-.418, p=0.645), social (Z=−0.221, 95% CI .399-.418, p=0.825), school (Z=−1.216, 95% CI .137-.151, p=0.224).

### Language and Communication

Descriptive statistics for total number of words used and understood by children are displayed in Table 4.

**Table 4.**
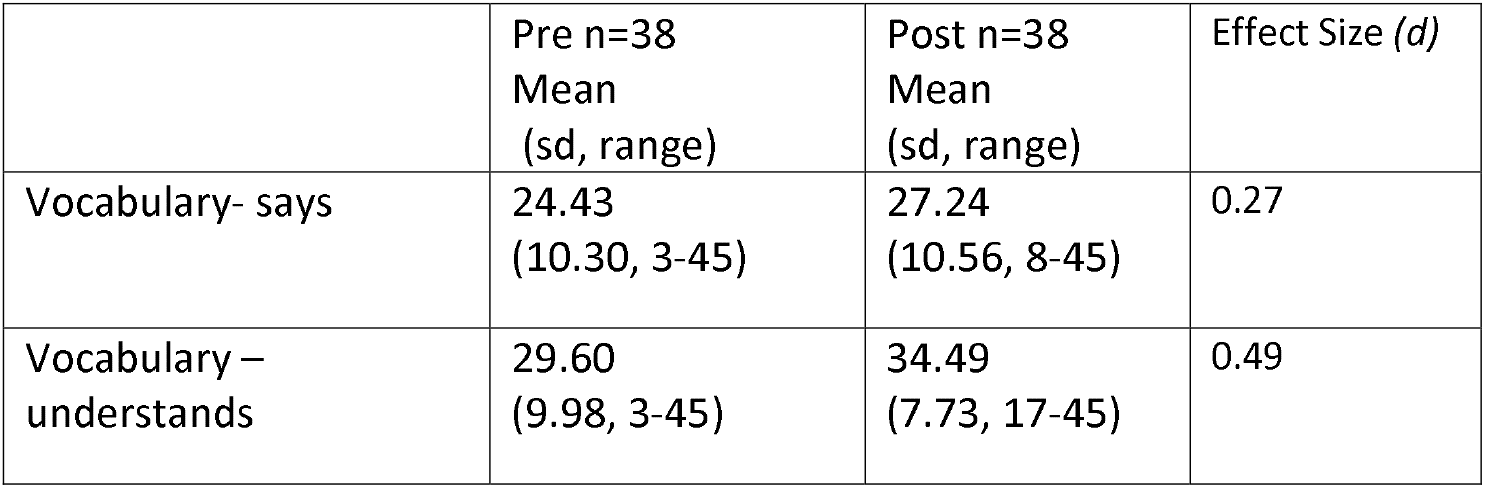
Descriptive statistics for pre- and post-expressive and receptive vocabulary scores.

|  | Pre n=38<br>Mean<br>(sd, range) | Post n=38<br>Mean<br>(sd, range) | Effect Size (d) |
| --- | --- | --- | --- |
| Vocabulary- says | 24.43<br>(10.30, 3-45) | 27.24<br>(10.56, 8-45) | 0.27 |
| Vocabulary – understands | 29.60<br>(9.98, 3-45) | 34.49<br>(7.73, 17-45) | 0.49 |

There was a significant effect of Zone West on the number of words children reported to understand (Z=−2.236, 95% CI .019-.025, p=0.025) with moderate effect size, and a small increase in the number of words children reported they used which was not significant (Z=0.684, 95% CI .488-508, p=0.494).

## DISCUSSION

The present study sought to provide early evidence for the impact of a novel SP programme embedded within education, ‘Zone West’, on the SEMH, quality of life and language skills of children aged 7-11 years old from deprived backgrounds with subclinical SEMH difficulties. Significant improvements were observed in children’s total SEMH difficulties, conduct difficulties and hyperactivity, their quality of life in terms of emotional and school functioning, and receptive vocabulary skills. Significant change observed by teachers and parents suggests children’s social-emotional presentation was observed both at home and within the school setting. Outcomes also suggest that the programme not only impacts SEMH, but also influences how children interact with the world around them; improving their ability to comprehend language input, manage emotions and function at school. However, no significant impacts on quality of life were reported by children themselves.

Moderate effect sizes found for SEMH and receptive vocabulary replicate those observed in educational research, above typical teacher effects and edging into the zone of desired effects for children^19^ and suggest these outcomes may be generalised beyond the current sample to the wider population. It is less clear whether outcomes observed for quality of life may be generalised due to small effect sizes. The ZW model and current outcomes however maintain recommendations for a personalised approach to healthcare delivery and facilitates calls for more effective partnerships between patients and professionals^10^. Given the formative nature of Zone West provision which highlights the importance of secure, consistent attachment relationships for mental health and wellbeing, it is hypothesised that relationships are underlying these outcomes. This hypothesis echoes existing evidence for social connectedness as a key mechanism underpinning SP effects^12^. The case for attachment being a key driver is further supported by the fact that despite LWs delivering targeted work with children to meet individual developmental needs as part of their provision, significant outcomes on broad measures are observed for the group as a whole.

Consistency of the LW-child relationship is maintained for all ZW children and this is supported by the school-based context of provision. The intensity of this relationship goes beyond adult models of SP, or more simple ‘active signposting’ approaches. In addition, improvement in receptive language as well as SEMH may evidence the close relationship between language difficulties and behaviour; that improvement in one domain supports improvement in the other. However the possibility of attachment as a key driver for change, and the possible influence of language and it’s relationship with SEMH require further exploration.

### Strengths and limitations

The study has provided proof of concept and quantitative evidence for education-based SP as a preventative strategy in child mental health which was lacking in existing SP models and literature. In addition, in a climate where SP for adults is growing in popularity, it has demonstrated the value of a SP approach for children and highlighted the potential value in implementing SP into education within an integrated care system. The study has also filled a gap in providing a defined model for the provision of SP for children and indicates the feasibility for teacher-referral as well as GP-referral.

The study has several limitations. It is limited in design due to a lack of control group; the inclusion of a comparative control or randomised control group given the high level of need of participants is challenging but it is recommended that this should be explored in the future. In addition, LWs led the collection of data with children which created a risk of reporting bias. This was implemented because it was collectively agreed amongst the team that this lay the foundations for, and strengthened, the LW-child relationship and engagement with intervention. The scoring and analysis of questionnaires however was completed by a researcher independent to data collection and intervention delivery. There was also substantial attrition in matched pre-post data which reduced study power, however despite this, sample size remained over the initial target of 70.

## Conclusions

Children’s poor mental health is a public health crisis that requires urgent up-stream prevention. The current model is presented as an approach that could offer support and relief to over-stretched primary and secondary care services and further work is required to explore direct impacts on mental health services. However, there may be a role for social prescribing embedded within education as a preventative strategy in children’s mental health and wellbeing, particularly for children at-risk due to the effects of poverty. Further work is required to assess whether the improvements observed are sustained long-term, the relative importance of relationships and attachment, asset attendance and theory that underpin these changes, and if the replicability of the model is possible outside of its current locality.

## Data Availability

All data produced in the present study are available upon reasonable request to the authors

## ACKNOWLEDGMENTS

We would like to thank all the primary schools, Primary Care Networks and families in West Newcastle, UK, who took part in Zone West, for their valuable commitment and contribution.

## FUNDING AND ETHICAL APPROVAL

The study was funded by the Northumberland, Tyne and Wear Community Foundation [grant number FR-000003976] and received ethical approval by Leicester South (REC reference:19/EM/0369 IRAS project ID: 266176) and was sponsored by Newcastle University (Reference 1824).

