## Supplemental material for "LINK WORKER LED SOCIAL PRESCRIBING AS EARLY INTERVENTION FOR CHILDREN WITH SOCIAL-EMOTIONAL MENTAL HEALTH DIFFICULTIES"

**Developmental Targets**

|  | A | B | C | D | Gauging  Progress |
| --- | --- | --- | --- | --- | --- |
|  | Understanding | Self-Awareness | Self-regulation/  Resilience | Agency, Mastery& Creativity |  |
| Learning &  School | Here’s what school says about how your work is going | What do you do when you find a piece of work difficult to complete? | Can you imagine being calm & engaged in front of tricky piece of work? | Do you get that feeling of ‘sitting on top’ of your schoolwork? | Teacher comment  Attendance  Attainment  Bloom’s Assessment |
| Physical Health  Exercise/Eating  Hygiene/Appearance | What does Health look like? | What does Health look like for you & what could you do to be more healthy? | What do you do to overcome setbacks and challenges to your health? | Can you map the help available & plan how to use it? | Parent comment  BMI  Asthma regulation  Absence for illness  GP Attendance |
| Emotional Development | Can you tell me what feelings are & when they happen? How does feeling x make people act? | What feelings do you have often? What would people see if you were feeling x, y or z? | What happens when you feel x? do you do something to help yourself along? | When a friend is having a hard time can you imagine what they are feeling? Is there a way of helping? | TSDQ  CSDQ  QoL |
| Social  Engagement | What is a social network and who is in it? | Who is in your network and what are the connections like (e.g. strong/week)? | When there is a problem in the network (and there often are) how do you repair bonds or make new ones? | Sometimes it seems that we have only a few friends and sometimes lots. How do you keep calm and keep looking outwards? | Parent/Asset comment  Asset Attendance  Self-reports |

**Zones of Development & Zones of Development: The How**

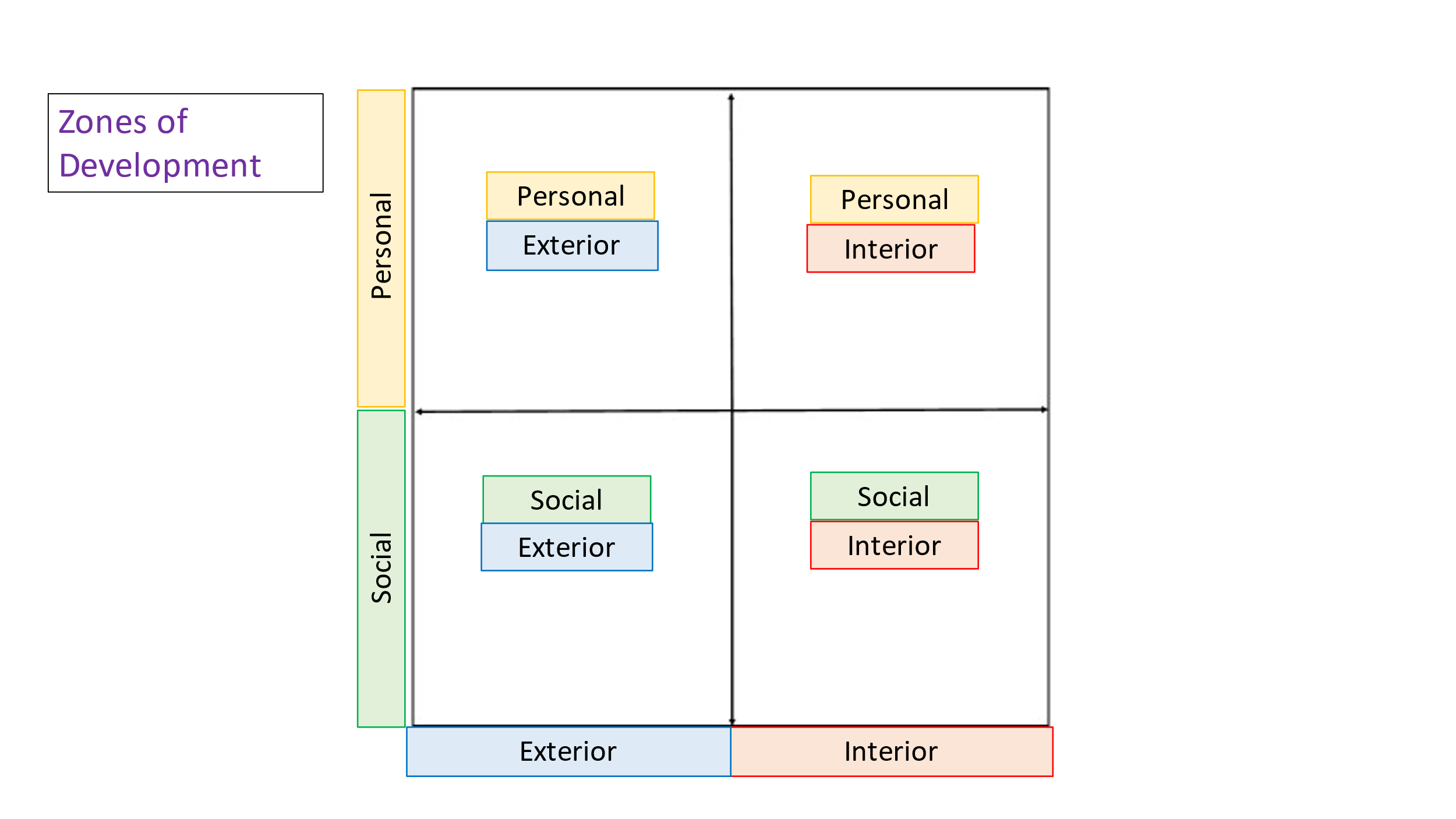

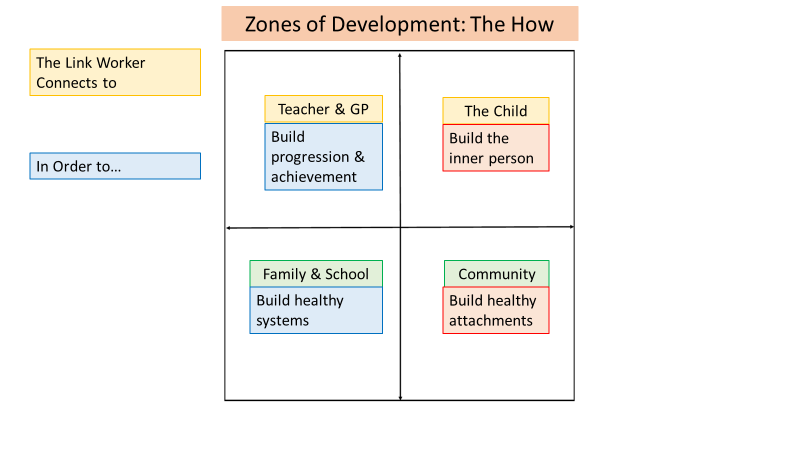
